# Acute kidney injury and death in severe falciparum malaria

**DOI:** 10.64898/2025.12.22.25342813

**Authors:** James A Watson, Andrea L Conroy, Anthony Batte, Ruth Namazzi, Michael T Hawkes, Kevin C Kain, Katherine Plewes, Stije Leopold, Hugh Kingston, Tran Tinh Hien, Chandy John, Nguyen Hoan Phu, Elizabeth C George, Ann Sarah Walker, Nicholas PJ Day, Thomas N Williams, Arjen M Dondorp, Kathryn Maitland, Nicholas J White, WWARN Severe Malaria Acute Kidney Injury Study Group

## Abstract

**Background:** Serum or plasma concentrations of creatinine and urea are the usual laboratory parameters measured to assess kidney function and disease severity in patients with suspected severe malaria. Creatinine is preferred for estimation of the glomerular filtration rate, but the current threshold for defining severe malaria (*>* 265 umol/L) is based on adult values and does not reflect the large difference between children and adults in normal values resulting from differences in muscle mass. We reevaluated these thresholds and their prognostic significance in severe malaria using individual patient data from large prospective studies.

**Methods:** We pooled individual patient data from studies of severe malaria. Patients were included in the primary analysis population if their age (if missing, imputed from weight or height), coma status on admission, and either an admission serum or plasma creatinine or urea measurement (blood urea nitrogen, BUN), were all recorded. The primary outcome was mortality by day 28. The secondary outcome was the admission plasma *Pf* HRP2 concentration (a measure of parasite biomass). Bayesian penalised spline regression models were used to characterise the age-specific relationships (interaction with age) between the outcome and the absolute creatinine, the creatinine fold change relative to the age/weight predicted baseline, and the absolute urea. The models adjusted for coma status and calendar year, with nested random additive effects by study site and study. We estimated threshold plasma or serum creatinine and urea concentrations associated with *>*5% mortality with probability *>* 0.9.

**Results:** We pooled individual patient data from 22,663 patients recruited into 14 studies of severe malaria of whom 16,441 were included in the primary analysis population. The majority (84%; 13,793/16,441) were children under 15 years of age. Current thresholds for renal impairment were associated with mortalities substantially greater than 5%, especially in children. A creatinine increase of more than three times the age/weight predicted baseline value was associated with 0.9 probability of mortality >5%. A urea of 10 mmol/L had approximately similar prognostic value. A urea *>*10 mmol/L was 3.5 times more common in adults than children. In children *<*15 years of age urea was a better prognostic indicator of death than creatinine and, unlike creatinine, correlated with the plasma *Pf* HRP2 concentration.

**Interpretation:** Admission serum or plasma creatinine *>*3 times the age/weight expected baseline value (KDIGO stage 3) can be used to define severe malaria. In children urea is a better predictor of the true infection biomass and a better prognostic indicator for death. We recommend the following as defining criteria for severe malaria; either the simple urea (BUN) threshold of 10 mmol/L for both children and adults, or a *>*3 times fold change in creatinine from predicted baseline.

## Background

Severe malaria is a medical emergency requiring rapid assessment and immediate treatment with artesunate, preferably by the parenteral route [1]. Severe falciparum malaria is characterised clinically by vital organ dysfunction usually as a direct consequence of a high parasite biomass and extensive parasitised erythrocyte sequestration. There is no absolute distinction between uncomplicated malaria and severe malaria, but rather a continuum of severity. The most appropriate definition of ‘severe malaria’ depends on the context. A more stringent definition is applied in a research setting compared to the inclusive indicators used in routine clinical care [1]. A general approach for defining severe malaria is to determine sets of clinical signs along with laboratory parameters which indicate an expected mortality greater than some threshold. For example, any clinical presentation with a diagnosis of malaria as the primary cause of illness and *>*5% chance of death could be categorised as ‘severe malaria’ [2]. That represents a mortality at least 50 times higher than that associated with ‘uncomplicated falciparum malaria’. This risk-based approach rationalises the definition, and allows it to be evidence based.

Current definitions of severe malaria used by the World Health Organisation (WHO) are based on expert consensus on the diagnostic and prognostic value of a range of clinical and laboratory parameters [1]. Acute kidney injury is an important manifestation of severe falciparum malaria. It is a common presentation in adults in low transmission settings [3], and for these patients effective renal replacement therapy reduces mortality substantially [4]. Clinically significant acute kidney injury was thought to be less common in children with severe malaria in high-transmission settings, although it has long been recognised that elevated biochemical measures of renal impairment were associated with increased mortality [5]. The renal impairment thresholds now used as criteria to define severe malaria (a plasma or serum creatinine *>*265 *µ*mol/L (3 mg/dL) or blood urea (or BUN) *>*20 mmol/L) were based on observational studies done in adults. They were not modified for children, in whom muscle mass, and thus baseline plasma or serum creatinine levels, are lower. Recent studies from East Africa indicate that renal impairment in children hospitalised with malaria is more common than appreciated previously because the current thresholds for determining renal impairment, based on adult normal values, are too high for children [6, 7]. Thus, the current creatinine threshold for severe malaria reflects a much greater reduction in glomerular filtration rate (GFR) in children, particularly the youngest children, than it does in adults. It is unclear what these high concentrations represent in terms of expected mortality in children presenting with suspected severe malaria.

We aimed to reevaluate and standardise the renal impairment thresholds to define severe malaria in adults and children by comparing the prognostic values of specific absolute and relative thresh-olds of serum creatinine and BUN using the largest pooled database comprising data from over 16,000 children and adults recruited into prospective studies of severe malaria. We compare the considered thresholds with the current thresholds for venous lactate concentration (one of the best measures of falciparum malaria disease severity which is independent of renal impairment [8, 9]). Together, this provides an evidence-based assessment of optimal thresholds to be used in clinical management to identify both children and adults with suspected severe malaria who are at a significantly elevated risk of death, and also to determine how general nephrology consensus criteria for acute kidney injury (e.g. KDIGO) perform in severe malaria.

## Methods

### Data

Randomised trials and prospective observational cohort studies enrolling adults or children with suspected severe malaria with at least one baseline BUN (or urea) or creatinine measurement were eligible for inclusion. Studies of children or adults with severe febrile illness not specifically recruiting severe malaria but which had a subset of patients with severe malaria (e.g. the FEAST trial which recruited children with shock and febrile illness [10]) were also eligible for inclusion. Studies were identified from the WHO 2014 review of severe malaria [1], and a systematic review of the literature of all studies done since 2014. For all identified studies, study authors were contacted and asked to upload the data to the secure IDDO data platform. All data were then curated to a standardised CDISC SDTM format prior to analysis. Extensive data checking was carried out before pooling of data.

### Analysis

The primary analysis population consisted of patients in the pooled database who had all of the following recorded:

1. A confirmed malaria infection at admission/enrolment (microscopy or rapid diagnostic test);
2. Age either directly recorded or imputed from weight or height;
3. Coma status at presentation;
4. Mortality outcome at hospital discharge or by day 28 after admission;
5. Admission plasma or serum creatinine or BUN measurement.

We use molar BUN concentrations which are the same as molar urea concentrations (whereas in mg/dL the blood urea is 2.14 times the BUN concentration).

The primary outcome was within-hospital mortality up to day 28. Many of the older studies did not systematically record the time to death and so we use a binary outcome of died versus survived. The secondary outcome was the plasma *Pf* HRP2 concentration: this was chosen because it is a proxy measurement of the parasite biomass and thus the severity of the falciparum malaria infection.

Three exposure variables were defined: a) the admission BUN; b) the admission creatinine; and the c) admission fold change in creatinine from the predicted baseline normal value. The primary analyses were performed on the complete case datasets: all patients who had a measured BUN or creatinine, respectively. All regression models included coma (yes/no; Glasgow Coma score <11 in adults, Blantyre Coma score <3 in children) as an independent predictor of outcome as this was recorded systematically across all studies and it is the strongest predictor of death. All regression models estimated age dependent relationships between the exposure variables and death (interaction term between age as a continuous variable and the exposure variable), modelled using a tensor-product smooth between the exposure variable and age with cubic regression splines. We also explored models that categorised age divided in three categories: *<* 5, [5, 15), and 15 years. Nested random intercepts were specified for each study and study site. A sensitivity analysis included a temporal spline for calendar year as there have been substantial improvements in treatment and supportive care over the 40 year period over which these studies were conducted.

For the analysis comparing the prognostic value of BUN versus creatinine, we included only the subset of patients who had both values recorded at baseline. For the comparative analysis examining the relationship between plasma lactate and mortality, we included only patients in the primary analysis population who also had a recorded baseline haemoglobin or haematocrit measurement. An interaction term was included for haemoglobin above 5 g/dL. If haemoglobin was missing, but the haematocrit was recorded, the haemoglobin was imputed using a 1/3 transformation from haematocrit (%) to haemoglobin (g/dL), see Supplementary Figure S9.

For the creatinine analysis, we compared the absolute creatinine values (*µ*mol/L) and the creatinine fold change relative to a predicted age and weight adjusted normal baseline value. To estimate the normal baseline values in children under 15 years we compared two separate approaches: the Pottel equation which is based on age only [11]; and the Schwartz equation which is based on height only. We then compared the predicted baseline creatinine values with the observed baseline values in a subset of patients recruited into the TRACT trial in Malawi who had follow-up creatinine and BUN both measured at 28, 90 and 180 days post admission (n=404 patients with ages ranging from 2 months to 12 years). This assumes the later measurements were equivalent to baseline values. The normal baseline values for patients over 15 years of age were estimated using the well established Cockroft-Gault equation, assuming a normal creatinine clearance of 100 mL/min. This equation was chosen as it incorporates both age and weight. Sensitivity analyses explored other predictive models for the predicted baseline creatinine, but they did not change the results.

We performed multiple imputation using chained equations for age, weight, height, mid-upper arm circumference and the key laboratory parameters (lactate, creatinine, BUN, bicarbonate, haemoglobin). The multiple imputation included the mortality outcome, sex, study and study site. We used random forests for the multiple imputation model as it allows for non-linear relation-ships between variables. Visual checks of the imputed versus complete case bivariate distributions were carried out to check multiple imputation outputs. Potassium values above 10 mmol/L were removed.

Bayesian generalised linear models with penalised spline terms were fitted for in-hospital mortality using the *brms* package which uses *stan* to estimate posterior distributions [12]. Mixed effects models were also fitted to the *Pf* HRP2 outcome using *lme4* (maximum likelihood estimation) [13]. All Bayesian models used default weakly informative priors to help computational convergence. Non-parametric estimates of proportions were done using conjugate Beta(1,1) priors. Euler diagrams (generalisation of Venn diagrams to up to 5 categories) were estimated from imputed data using the R package *eulerr* [14].

### Data and code

All R code can be found at https://github.com/Infectious-Diseases-Data-Observatory/severe-malaria-renal-failure. Pseudonymised participant data used in this analysis are available for access via the WWARN website (https://www.iddo.org/wwarn/accessing-data). Requests for access will be reviewed by a data access committee to ensure that use of data protects the interests of the participants and researchers according to the terms of ethics approvals and the principles of equitable data sharing. Requests can be submitted by email to via the data access form. WWARN is registered with the Registry of Research Data Repositories.

## Results

### Participants

Individual patient data from 14 studies were pooled, providing data from a total of 22,663 patients. After excluding one study with unreliable creatinine measurements and one study that had no urea or creatinine data, 16,441 patients from 12 studies had a recorded or imputed age, coma status, parasitological confirmation of malaria at baseline, a recorded mortality outcome, and at least one baseline measurement of creatinine or urea (BUN). These 16,441 patients were included in the analysis population (9,848 patients had creatinine measured on admission; 12,537 patients had urea (BUN) measured on admission; and 5,944 patients had both), Figure S1. Table 1 shows the demographic characteristics of the patients in the analysis population stratified by study. The majority of patients were children (n=13,793 *<*15 years) and from Africa (n=13,533). 10.4% of all patients died within 28 days of admission (n=1,715/16,441).

**Table 1:** Summary characteristics of the patients included in the pooled meta-analysis, by clinical study. The top set of studies were in Asia; the bottom set of studies were in Africa.

| Study | Year | N | Age (median, IQR) | Mortality (n, %) | Countries | Creatinine measured | BUN measured | Lactate measured |
| --- | --- | --- | --- | --- | --- | --- | --- | --- |
| Thailand cohort | 1980-2006 | 588 | 25 [19-35] | 18.0 | Thailand | 485 | 579 | 346 |
| AQ Vietnam [15] | 1991-1996 | 558 | 30 [22-40] | 14.9 | Vietnam | 558 | 260 | 477 |
| Dong Nai [16] | 1992-1995 | 96 | 6 [3-9] | 8.3 | Vietnam | 75 | 83 | 57 |
| AAV [17] | 1996-2003 | 355 | 35 [24-43] | 9.6 | Vietnam | 354 | 303 | 320 |
| SEAQUAMAT [18] | 2003-2005 | 1,311 | 25 [18-36] | 18.2 | Bangladesh, Myanmar, India, Indonesia | 0 | 1,311 | 0 |
| Kilifi cohort [19] | 1995-2023 | 3,472 | 3 [2-5] | 10.1 | Kenya | 3472 | 0 | 0 |
| AQUAMAT [20] | 2005-2010 | 4,389 | 3 [2-4] | 10.0 | Tanzania, Democratic Republic of the Congo, Uganda, Mozambique, Ghana, Rwanda, Kenya, Gambia, Nigeria | 0 | 4,389 | 0 |
| Conroy 2019 [21] | 2008-2015 | 495 | 3 [2-5] | 7.1 | Uganda | 479 | 492 | 460 |
| FEAST [10] | 2009-2011 | 1,353 | 2 [1-3] | 10.4 | Uganda, Kenya, Tanzania | 608 | 1,353 | 1,292 |
| NO Uganda [22] | 2011-2013 | 165 | 2 [1-3] | 8.5 | Uganda | 163 | 119 | 161 |
| Namazzi 2022 [23] | 2014-2017 | 594 | 2 [1-3] | 7.1 | Uganda | 594 | 592 | 569 |
| TRACT [24, 25] | 2014-2017 | 3,065 | 3 [2-5] | 7.3 | Uganda, Malawi | 3,060 | 3,056 | 3,031 |

### Age and weight dependent distribution of creatinine and urea in severe malaria patients

Figure 1 shows the relationship between weight (kg, square root scale) or age (years, square root scale) and the admission serum or plasma creatinine (left panels) and urea (right panels; both on the log_10_ scale). For both creatinine and urea there was, as expected, a clear relationship between weight/age and admission values, with substantial inter-individual variation. When comparing children (defined as patients younger than 15 years of age) versus adults (≥15 years), the median serum or plasma creatinine values were 45 *µ*mol/L (IQR: 31-64) versus 155 *µ*mol/L (IQR: 113-256), respectively; i.e. they were approximately 3.4 times higher in adults. For urea the median values were 4.6 mmol/L (IQR: 2.9-7.5) versus 10.9 mmol/L (IQR: 6.4-21.4), i.e. approximately 2.4 times higher in adults. The larger difference between creatinine values in adults versus children likely reflects the stronger dependence of creatinine on muscle mass and hence weight/age. This was also reflected by a systematic difference in the creatinine/urea ratios between children and adults (Figure 2). The relationship between weight and urea was consistent across five large studies [10, 18, 20, 21, 23] which all used the portable i-stat machine (Abbott) to measure BUN (Supplementary Figure S2).

**Figure 1:**
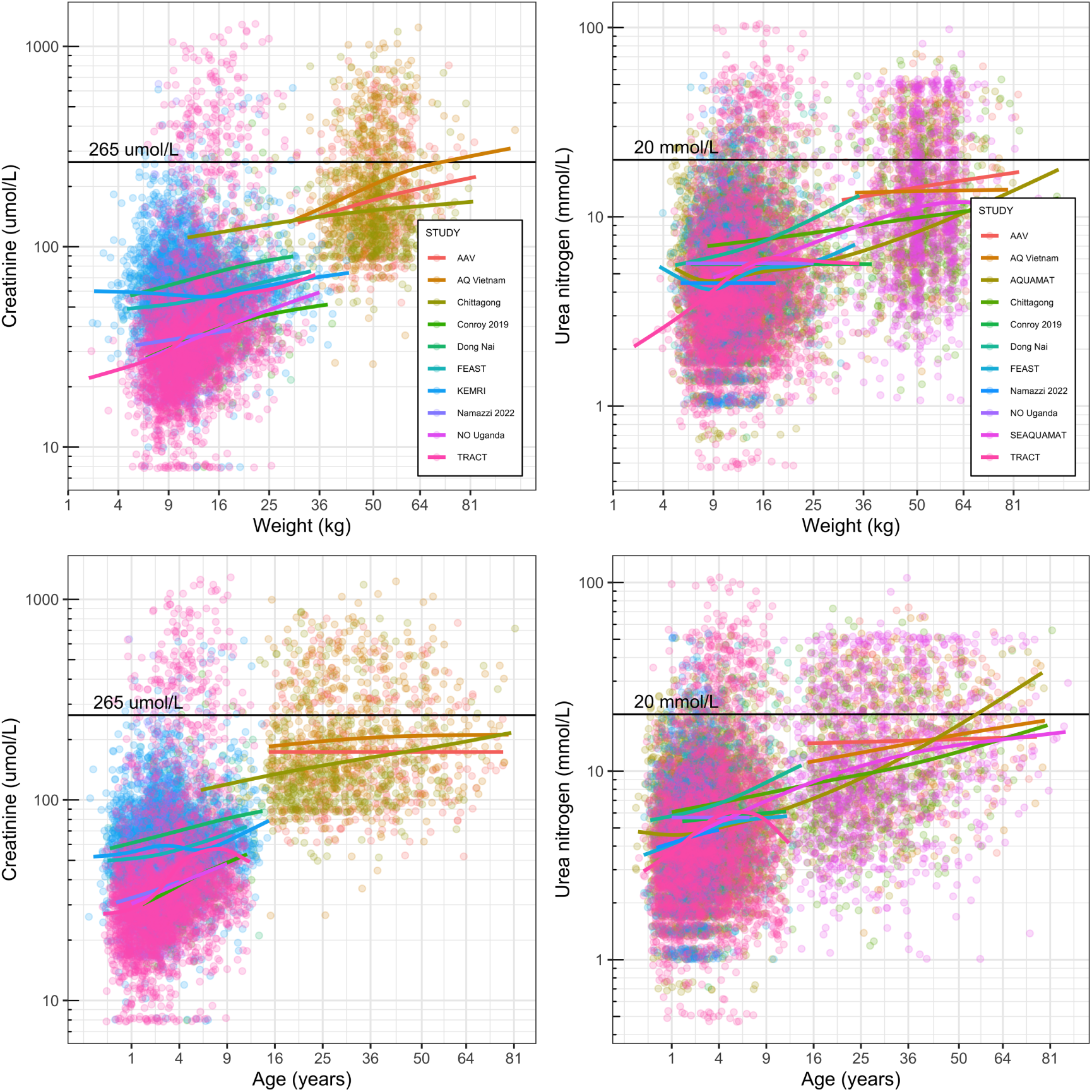
Relationship between weight (top panels) or age (bottom panels; both on the square root scale) and plasma or serum concentrations of creatinine (left panels) and urea (right panels, both on the log_10_ scale). Lines show spline regression fits by study. Horizontal lines show the currently used severe malaria definition thresholds of 265 *µ*mol/L for creatinine and 20 mmol/L for urea.

**Figure 2:**
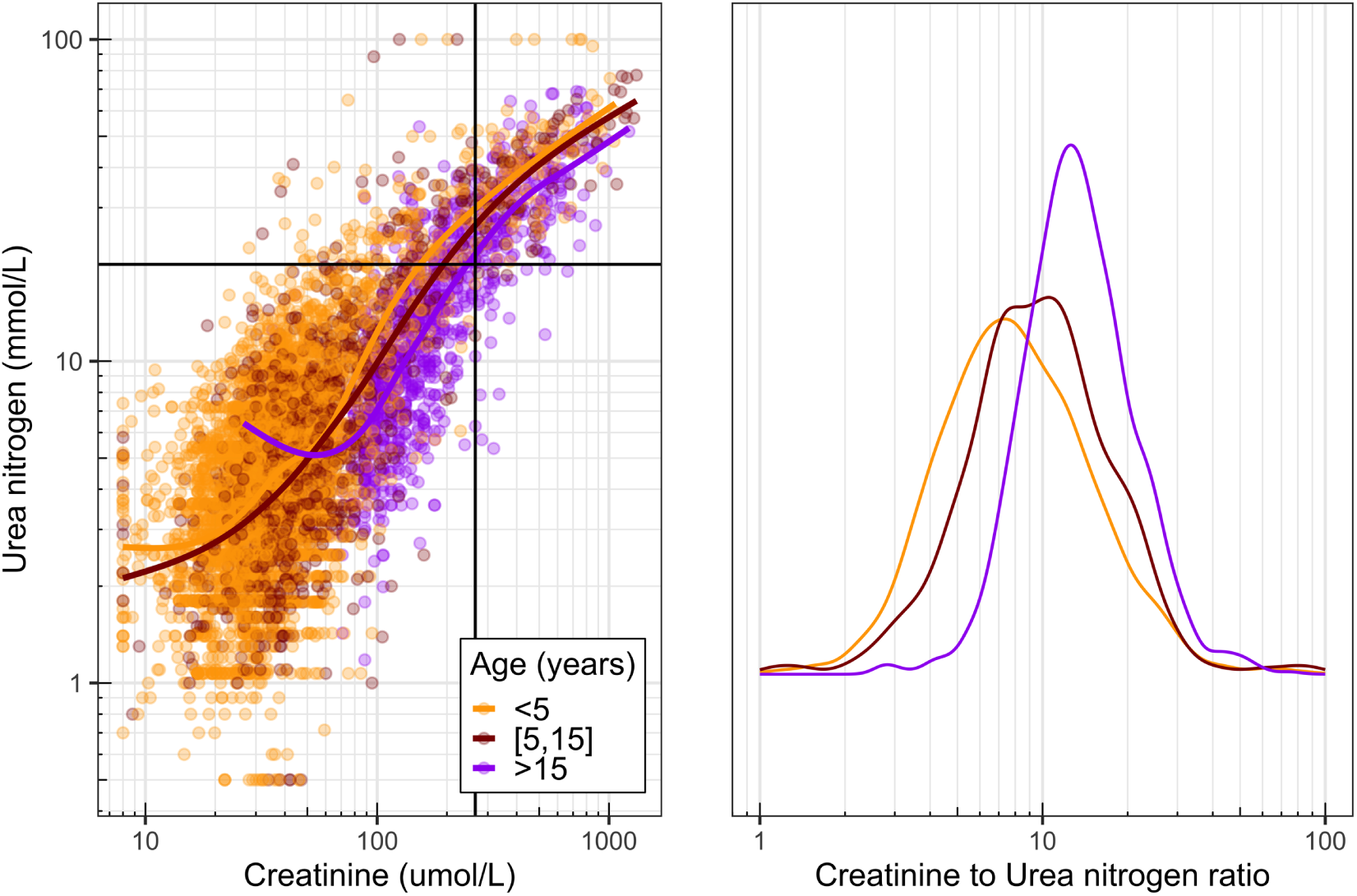
Relationship between plasma or serum creatinine and urea concentrations in patients in whom both were measured on admission (*n*=5,944) stratified by age category. In the left panel the vertical and horizontal thick lines show the current creatinine (265 *µ*mol/L) and urea (20 mmol/L) thresholds used to define severe malaria. The right panel shows the distribution of ratios stratified by age category.

In children under 15 years of age, only 2.4% had a plasma or serum creatinine above 265 *µ*mol/L (n=205/8473); whereas in older children and adults, 24.4% had a creatinine above 265 *µ*mol/L (336/1375). For urea, 4.5% of patients younger than 15 years of age had an admission value above the current threshold of 20 mmol/L (n=463/10248), versus 27.2% for patients older than 15 years (n=622/2289). The aggregate day 28 case fatality ratios were 8.1% in children younger than 15 years (n=1,116/13,793) and 17.1% in those older than 15 years (n=453/2,648).

### Prognostic value of creatinine and BUN

To explore the relationships between creatinine, urea and mortality by day 28 we divided patients into four categories for renal biomarkers (creatinine: ≤65, (65-130], (130, 265], *>*265 *µ*mol/L; urea: ≤5, (5-10], (10, 20], *>*20 mmol/L). As coma on admission has the greatest prognostic significance of all clinical variables, we estimated the proportion of patients who died in each creatinine and urea category, subdivided by coma status on admission (yes/no) and age (*<*15 years versus ≥15 years). For patients meeting the current WHO renal impairment threshold for defining severe malaria (plasma or serum creatinine *>*265 *µ*mol/L or urea *>*20 mmol/L), the observed mortality was considerably greater than 5% in both children and adults, even without coma on admission (all confidence intervals excluded mortality less than 10%), Figure 3. Mortality was greater than 5% for threshold values of 130 *µ*mol/L for creatinine and 10 mmol/L for urea (i.e. half the current thresholds).

**Figure 3:**
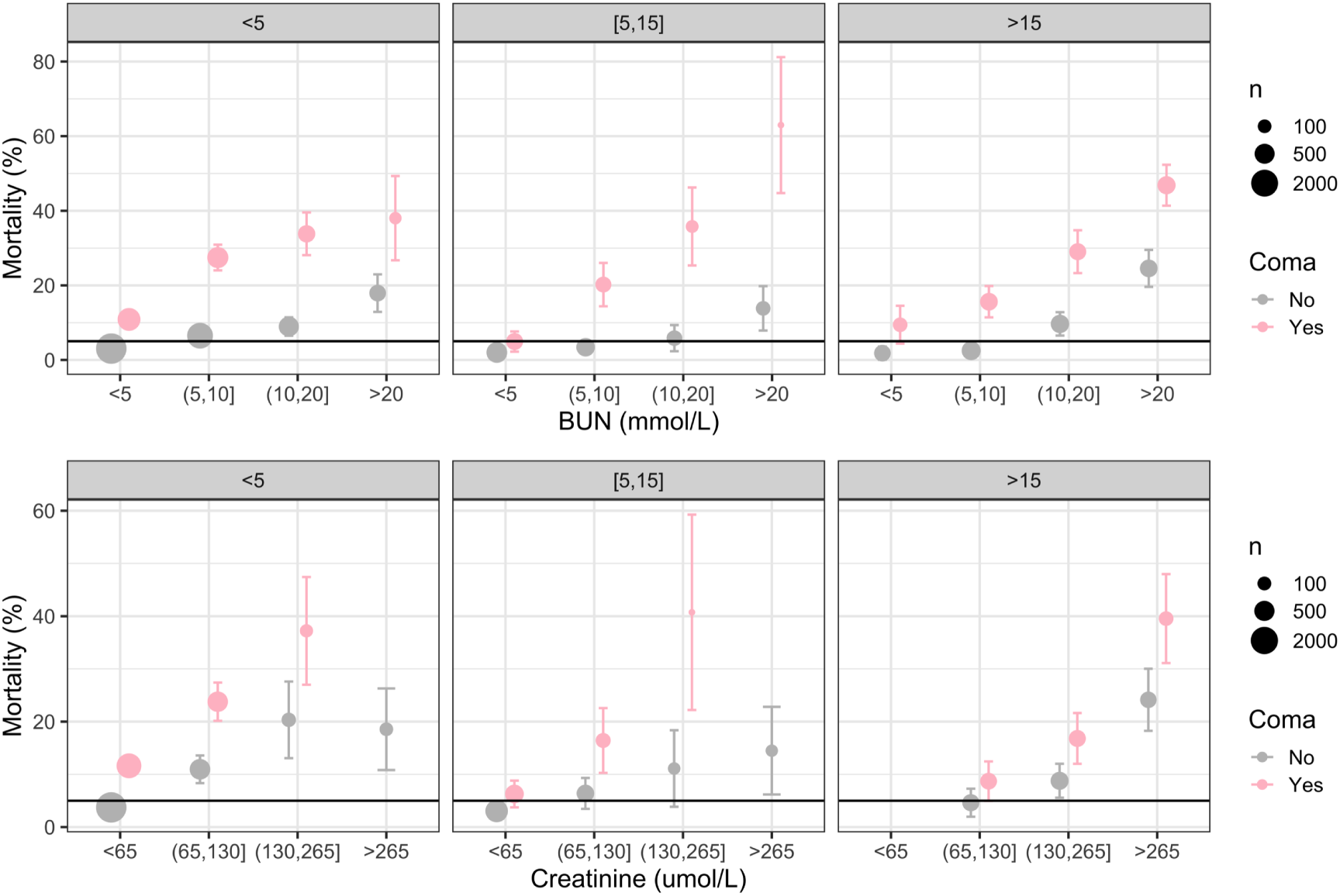
Observed mortality in patients with suspected severe malaria, stratified by coma status and age. The horizontal line shows a 5% mortality threshold. Points shows mean observed mortality with 95% uncertainty intervals derived from a Beta(1,1) prior. The size of the points is proportional to √*n*. Categories with fewer than 20 datapoints were discarded.

There was clear heterogeneity between the absolute plasma or serum creatinine value and mean case fatality ratio by age (Figure 3). The same absolute creatinine thresholds implied substantially different expected mortalities in children versus adults. This was less apparent for urea. In order to harmonise comparisons between adults and children, we estimated a creatinine fold change from predicted baseline values and used that as the primary exposure variable. Using 6-month follow-up data from children in Malawi recruited into the TRACT trial to provide normal paediatric baseline values, there was a clear linear relationship between age and baseline creatinine, the slope of which was well approximated by the Schwartz equation (it had a systematic upwards bias but the residuals were less correlated with the baseline value relative to the Pottel equation, Figure 4 and Supplementary Figure S3). Height, age and weight all had approximately the same predictive value (explaining roughly 50% of the variance in baseline creatinine values in these data). As expected, urea did not vary by age or weight, apart from in the youngest children (*<*2 years) (Figure 4). Very young children (under 1 years of age) had baseline urea concentration around 30% lower than the older children (approximately 2.0 mmol/L versus 2.85 mmol/L).

**Figure 4:**
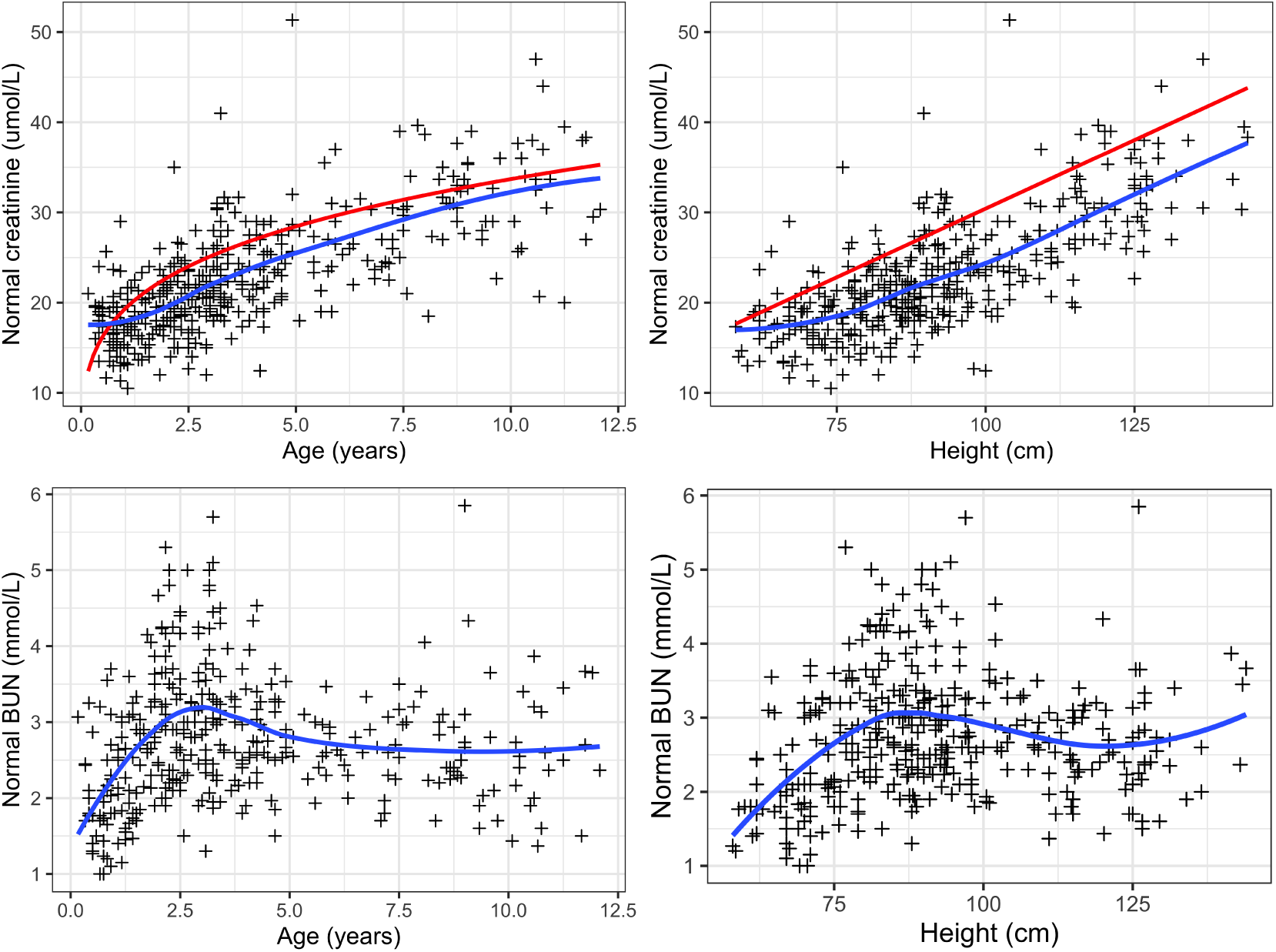
Postillness creatinine and urea (BUN) concentrations from 404 Malawian children enrolled into the TRACT trial (n=404 had postillness creatinines and n=361 had postillness urea). The blue lines show spline fits and the red lines show either the Schwartz equation prediction (height versus creatinine) or the Pottel equation prediction (age versus creatinine).

As expected, given the large variation in baseline creatinine by age, there was considerable variation in predicted mortality as a function of the absolute creatinine values by age (Figure 5). In patients without coma, the probability of *>*5% mortality exceeded 90% for a creatinine around 80 *µ*mol/L in children, compared with around 200 *µ*mol/L in adults. However, under the same model structure, there was very little residual heterogeneity by age in the predicted mortality relationship when using the creatinine fold change (this was not sensitive to the exact method used for the predicted baseline creatinine). A three times increase in predicted baseline creatinine (i.e. KDIGO stage 3) was associated with a mortality >5% in all age groups (Figure 5). For urea, there was considerably less heterogeneity between the absolute value and the predicted risk of death. Older children and adults had slightly higher threshold values (between 12 and 14 mmol/L) associated with >0.9 probability that death would occur in *>*5% of patients, Figure 5.

**Figure 5:**
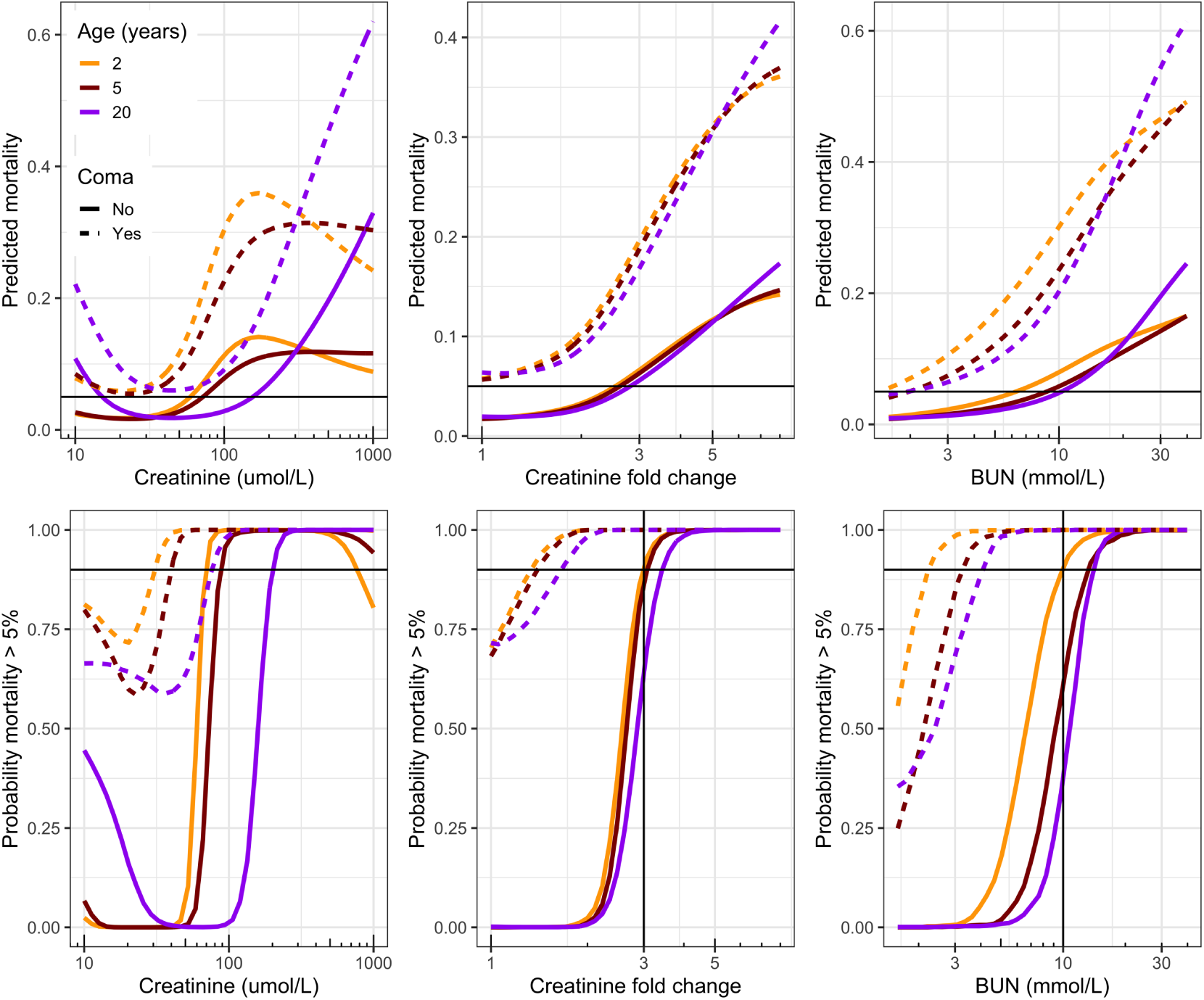
Modelled relationship between mortality and the fold change from baseline for creatinine (left) or urea (BUN) (right) in children and adults diagnosed with severe malaria. Top panels show the mean predicted mortality as a function of creatinine fold change or urea, stratified by age group and coma status; the bottom panels show the posterior probability that the mortality is greater than 5%.

Overall a plasma or serum urea (BUN) *>*10 mmol/L had a very similar prognostic value to a plasma lactate *>*5 mmol/L. In a multivariable logistic regression model which included age as a continuous variable and coma status, the adjusted odds-ratio for death was 2.9 (95% CI: 2.3-3.5) for urea *>*10 mmol/L and 2.7 (95% CI: 2.2-3.4) for lactate *>*5 mmol/L (n=5,718 patients had both lactate and urea measured). In comparison, in the same model, the adjusted odds ratio for death in those with coma on admission was 4.1 (95% CI: 3.3-5.2).

In a multivariable logistic regression model comparing creatinine fold change and urea (BUN) (both on the log_2_ scale), with an interaction by age category, adjusted for coma status, a doubling in urea was a better predictor of death than a doubling in creatinine fold change in all age groups under 15 years. In adults urea and creatinine had similar prognostic value (Supplementary Figure S4 and S5). A doubling in plasma or serum creatinine relative to the age/weight predicted baseline was associated with an adjusted odds ratio for death of 2.4 (95% CI: 1.3-4.5) in patients *>*15 years, whereas for all age groups in children under 15 years the credible intervals crossed 1. In comparison, a doubling of urea was associated with an increased risk of death in all age groups with estimated odds ratio between 1.5 (*>*15 years) and 2.3 (children between 5 and 15 years).

### Hyperkalaemia in severe malaria

Hyperkalaemia is associated with both acute kidney injury and acidosis (of any cause). Clinically significant hyperkalaemia (*K*^+^ *>*6.5 mmol/L) on admission was recorded in 3.9% (583/14,952) of patients with admission potassium values. Nearly all were in children (only 19 out of 2,377 [0.8%] were in adults). Elevated creatinine or urea concentrations were only weakly predictive of hyperkalaemia (Figure 6) with a large number of children with hyperkalaemia and BUN *<*10 mmol/L. Very low bicarbonate (*<*10 mEq/L) was the strongest predictor of hyperkalaemia.

**Figure 6:**
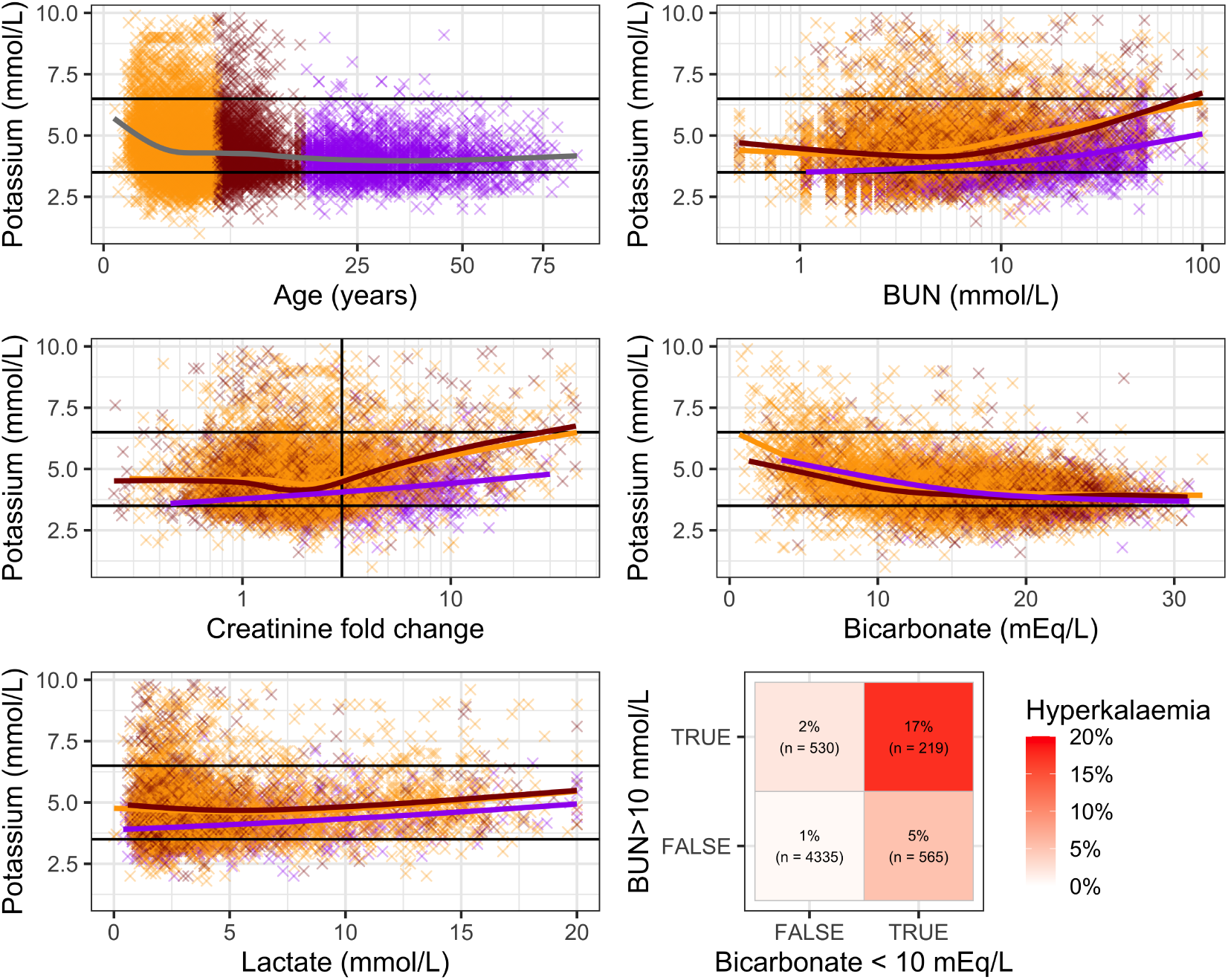
Admission potassium as a function of age, creatinine fold change from predicted baseline, BUN, and lactate. Colours correspond to age groupings (<5, [5-15), 15.) The horizontal lines show thresholds of 6.5 mmol/L (hyperkalaemia) and 3.5 mmol/L (hypokalaemia).

### Association of renal impairment with parasite biomass

In the pooled database, there were 4,116 patients from 4 studies with admission plasma *Pf* HRP2 and creatinine both measured, and 7,697 patients from 5 studies with admission plasma *Pf* HRP2 and urea both measured. All were children under 15 years of age. Plasma *Pf* HRP2 is the best quantitative marker of the total parasite biomass and thus the true underlying severity of the infection [26]. We assessed its relationship with creatinine fold change and urea. In the TRACT trial, there was a large subgroup of patients with very low plasma *Pf* HRP2 and high creatinine and high BUN with an unexpectedly low case fatality ratio (Supplementary Figures S6 and S7). The reason for this was unclear. In patients with a plasma *Pf* HRP2 >1,000 ng/ml (i.e. patients likely to have true severe malaria [27]), there was little correlation between plasma *Pf* HRP2 and creatinine fold change, but there was a strong correlation between plasma *Pf* HRP2 and urea (BUN) (Supplementary Figure S8).

### Attributable fraction of acute kidney injury in children and adults

We used the full analysis dataset with multiple imputation for missing data to estimate the proportion of patients with a presentation of renal impairment (urea >10 mmol/L), versus cerebral malaria, severe anaemia (haemoglobin *<*5 g/dL), and lactic acidosis (lactate *>*5 mmol/L). 75% of patients in the analysis dataset (12,357/16,441) met one of these categories (10,134/13,793 children and 2,119/2,648 adults). Figure 7 shows the estimated individual proportions and overlaps for each category. A urea >10 mmol/L was a dominant manifestation in adults (54%), but only occurred in 16% of children (i.e. 3.5 times more common in adults). Very high urea concentrations (*>*30 mmol/L) occurred in approximately 3.5% of children and 15% of adults (i.e 4 times more common in adults). Analysis of the complete case data gave very similar proportions.

**Figure 7:**
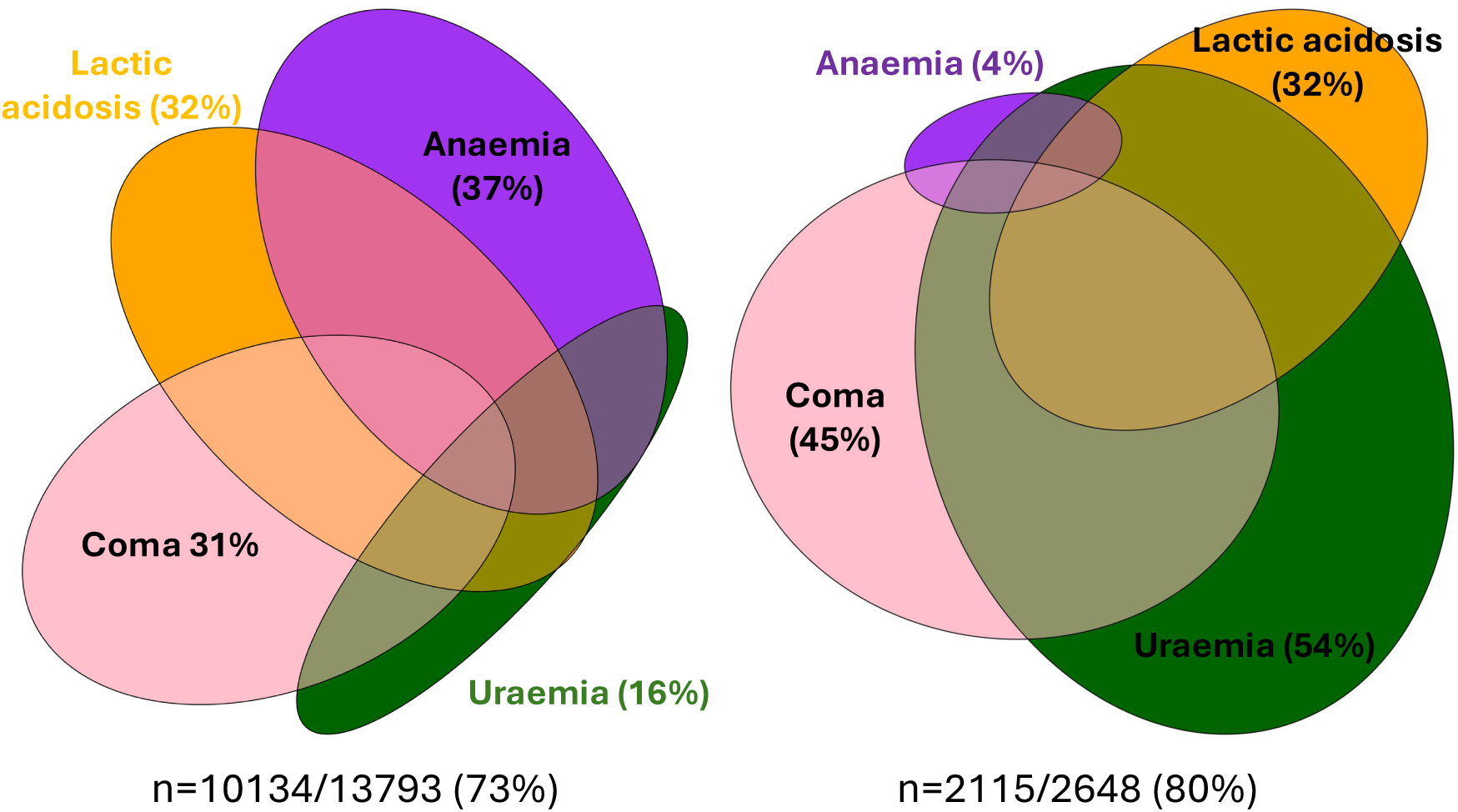
Euler diagrams showing the proportions of patients (left: children <15 years; right: adults) meeting the severe malaria criteria for each of the 4 major organ system involvements (Anaemia: haemoglobin<5 g/dL for both children and adults; uraemia: urea *>*10 mmol/L; lactic acidosis: lactate*>*5 mmol/L). Note not all combinations can be shown for 4 categories. Using the imputed BUN and lactate values, 73% of children and 80% of adults in the dataset met one of these 4 categories.

## Discussion

Severe falciparum malaria is a multi-system disease and a major cause of death in endemic areas. Identifying patients with severe malaria is essential for appropriate triage, and also for the correct evaluation of therapies. The current definition of severe malaria used in clinical research comprises a list of clinical or laboratory abnormalities, any one of which fulfils the definition. All, except the anaemia criterion (haemoglobin less than 5 g/dL), are associated with a mortality *>*5% [2]. Renal impairment (acute kidney injury) is an important manifestation of severe malaria, but the biochemical thresholds used to define it have not been calibrated to a mortality threshold of 5%, and have been based on values three times the upper limit of normal in adults, whereas most deaths from severe falciparum malaria occur in children. The degree of renal impairment is usually assessed from a plasma or serum measurement of creatinine or urea. Plasma or serum creatinine is more accurate than urea as an indirect measure of the glomerular filtration rate (GFR) but, as the level depends on the muscle mass, values in children are significantly lower than those in adults, particularly if they are malnourished. Creatinine levels should therefore be adjusted for lean body mass to provide comparable estimates of GFR. As a consequence the current severe malaria threshold (*>*265 *µ*mol/L) is an insensitive indicator in children.

This individual patient analysis of the largest data set of adults and children diagnosed with severe malaria derived from prospective studies in research centres confirms that the admission fold-change in plasma or serum creatinine (above the predicted normal value) is highly predictive of death and that a three-fold increase is associated with a mortality above 5%. This corresponds to a KDIGO stage 3 classification. Using age or weight adjustment the normal values are satisfactorily predicted by widely used equations (e.g. the Schwartz equation, Figure 4). These age/weight corrections removed nearly all the observed heterogeneity between creatinine and mortality outcomes across age groups. Age based thresholds suitable for African children have been proposed [7]. As expected creatinine and urea levels were strongly correlated. In severe malaria increased metabolism and variable dehydration both contribute to the rise in urea. However the urea concentration (often reported as BUN) was a better prognosticator than the creatinine concentration in children. In addition, apart from in very young children, urea was not strongly determined by weight or age (Figure 4) and, unlike creatinine, there was not large heterogeneity in its predictive value. Across the age range a urea over 10 mmol/L was associated with a risk of death of 5% or more. A urea threshold of 10 mmol/L has similar prognostic value as a lactate threshold of 5 mmol/L.

Severe falciparum malaria is associated with a large sequestered parasite biomass. This is reflected in high plasma concentrations of *Pf* HRP2 (a major parasite protein). This helps to separate children with true severe malaria from those with a severe febrile illness (often sepsis), and incidental parasitaemia. In children with true severe malaria, increases in urea but not creatinine concentrations were predictive of the parasite biomass. Severe malaria is overdiagnosed in children in endemic areas. It has been estimated that approximately one third of African children diagnosed as severe malaria do not have it – with sepsis being the most likely alternative [27]. Sepsis is a major cause of AKI in low resource settings, both in malaria endemic and non-endemic regions. Renal impairment in sepsis is associated with a worse prognosis, slower recovery and may require renal replacement therapies. Nevertheless, while a substantial proportion of children in this series may not have had true severe malaria (particularly those with low plasma *Pf* HRP2 and high platelet counts), this is unlikely to have affected the prognostic significance of elevated urea or creatinine concentrations.

In most patients urea concentrations were well below the range associated causally with uraemic complications. Renal impairment contributes to metabolic acidosis which is a major cause of death in severe malaria. Lactic acid and gut derived acids are the other main contributors. Acidosis is measured as reduction in the standard bicarbonate (sHCO3). Adjusting for lactate the serum or plasma concentrations of creatinine and BUN remained highly significant correlates of the sHCO3, reflecting the direct contribution of renal impairment to acidosis. Of the four main indicators for renal replacement therapies (uraemia, fluid overload with oliguria, acidosis and hyperkalaemia), the plasma potassium had the greatest variability in its association with renal impairment. Hyperkalaemia in children was recorded across the range of urea or creatinine concentrations and only at the very highest concentrations of urea or creatinine was there evidence of a consistent elevation in plasma potassium. Thus renal impairment was not the main driver of hyperkalaemia. In clinical practice most hyperkalaemia is spurious, resulting from blood sample haemolysis. In this large series high potassium values were most frequently observed in younger children (age <2 years) (Figure 6). Severe acidosis is associated directly with hyperkalaemia. The extreme acidosis and associated hyperkalaemia observed in a subgroup of children aged < two years, which was largely independent of renal impairment, is notable.

This individual patient data meta-analysis confirms the important prognostic value of measures of acute kidney injury in both children and adults diagnosed with severe malaria. Whether sepsis associated AKI is associated with a more severe degree of renal impairment than severe malaria in hospitalised children warrants further study. Measurement of urea (expressed either as urea or urea nitrogen) concentration is widely available and inexpensive, does not require adjustment for muscle mass and is less susceptible to measurement errors than creatinine. A value of 10 mmol/L confers a 5% or higher risk of death across all age groups.

## Data Availability

Pseudonymised participant data used in this analysis are available for access via the WWARN website (https://www.iddo.org/wwarn/accessing-data). Requests for access will be reviewed by a data access committee to ensure that use of data protects the interests of the participants and researchers according to the terms of ethics approvals and the principles of equitable data sharing. Requests can be submitted by email to via the data access form.

## Acknowledgements

We thank all participating patients and everyone involved in the original studies. NJW is a Principal Research Fellow funded by the Wellcome Trust (093956/Z/10/C). JAW is a Sir Henry Dale Fellow funded by the Wellcome Trust (223253/Z/21/Z). TNW was supported by a Senior Research Fellowship from the Wellcome Trust (202800/Z/16/Z). ASW and ECG are supported by core support to the MRC Clinical Trials Unit at UCL [MC_UU_00004/05]. ASW is an NIHR Senior Investigator. Funding from Wellcome Collaborative Grant (SMAART: Grant Number 209265/Z/17/Z) provided support for additional sample analysis from the FEAST biobank.

## Authorship and contributions

WWARN Severe Malaria Renal Failure Study Group: James A Watson, Andrea L Conroy, Anthony Batte, Ruth Namazzi, Michael T Hawkes, Kevin C Kain, Katherine Plewes, Stije Leopold, Hugh Kingston, Tran Tinh Hien, Chandy John, Nguyen Hoan Phu, Elizabeth C George, Ann Sarah Walker, Nicholas PJ Day, Thomas N Williams, Arjen M Dondorp, Kathryn Maitland, Nicholas J White.

*Infectious Diseases Data Observatory, Oxford, UK*: James A Watson. *Center for Global health and Tropical Medicine, University of Oxford*: James A Watson, Stije Leopold, Nguyen Hoan Phu, Nicholas PJ Day, Arjen M Dondorp, Nicholas J White. *Indiana University School of Medicine*: Andrea L Conroy, Chandy John. *Makerere University College of Health Sciences*: Anthony Batte, Ruth Namazzi. *Mahidol Oxford Tropical Medicine Research Unit, Bangkok, Thailand*: Katherine Plewes, Stije Leopold, Hugh Kingston, Nicholas PJ Day, Arjen M Dondorp, Nicholas J White. *Oxford University Clinical Research Unit, Vietnam*: Tranh Tinh Hien, Nguyen Hoan Phu *University of British Columbia, Vancouver, Canada*: Michael T Hawkes. *University of Toronto, Toronto, Canada*: Kevin C Kain. *KEMRI Wellcome Trust Research Programme, Kilifi, Kenya*: Thomas N Williams, Kathryn Maitland. *Institute Of Global Health and Innovation, Imperial College, London*: Thomas N Williams, Kathryn Maitland. *MRC Clinical Trials Unit at UCL, London*: Elizabeth C George, Ann Sarah Walker.

## Conflict of Interest Disclosures

The authors declare no conflict of interest.

## Supplementary Figures

**Figure S1:**
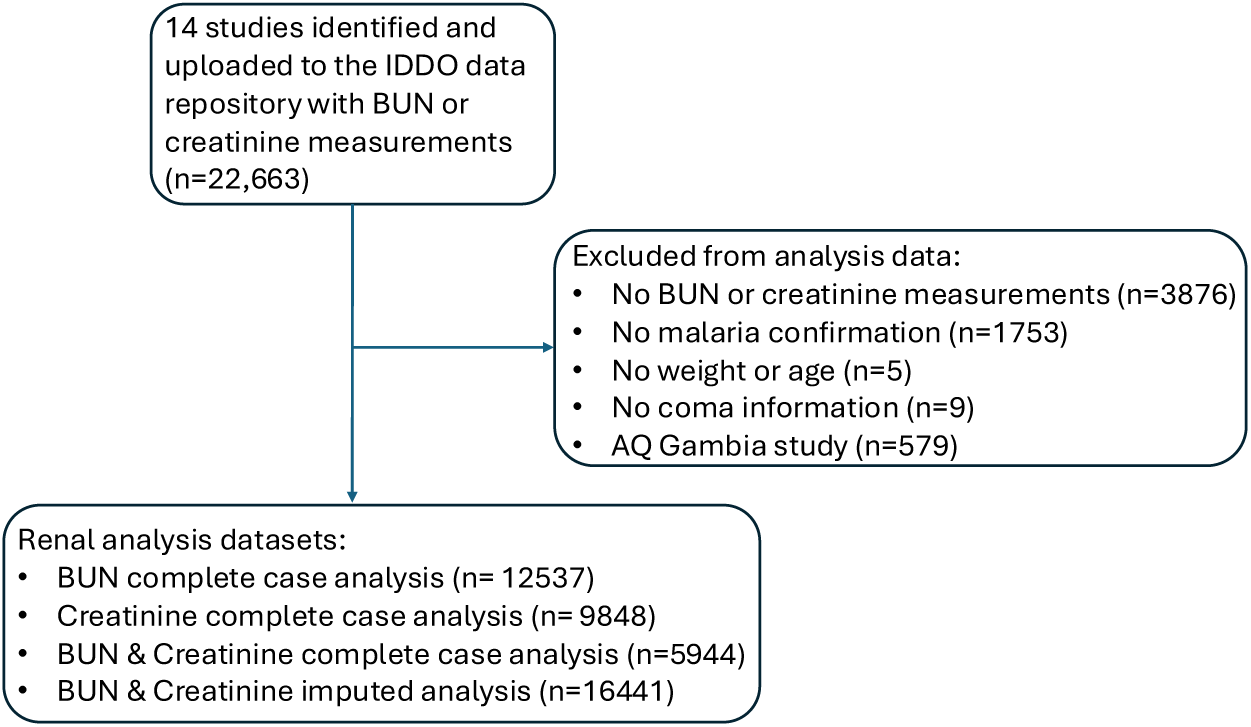
Analysis flow for the inclusion of data into the primary analysis population.

**Figure S2:**
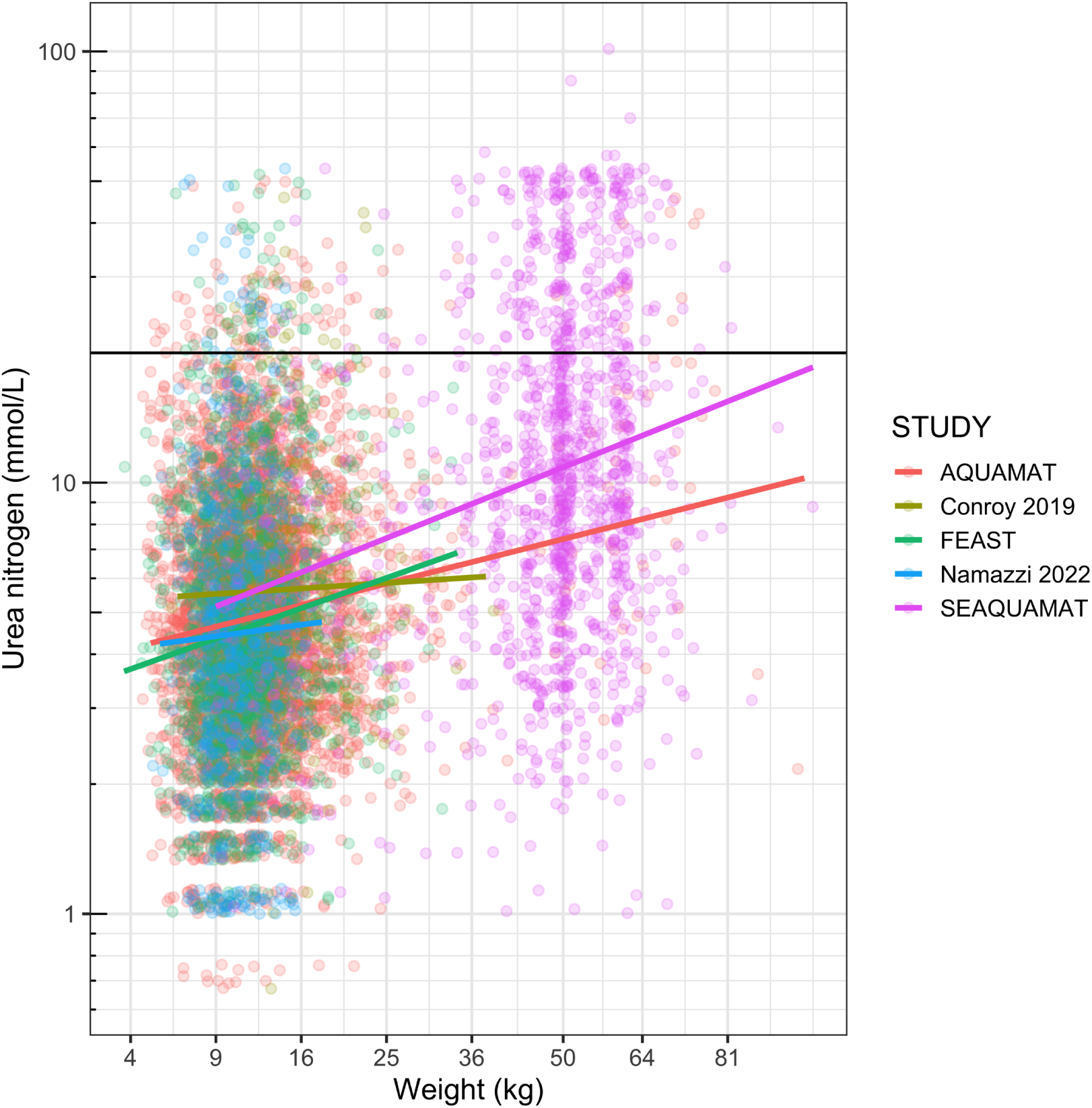
Relationship between weight (square root scale) and BUN (mmol/L) for 5 large studies [10, 18, 20, 21, 23] which all used the i-stat machine to measure BUN (n=8,137 patients). Linear regression lines by study are shown.

**Figure S3:**
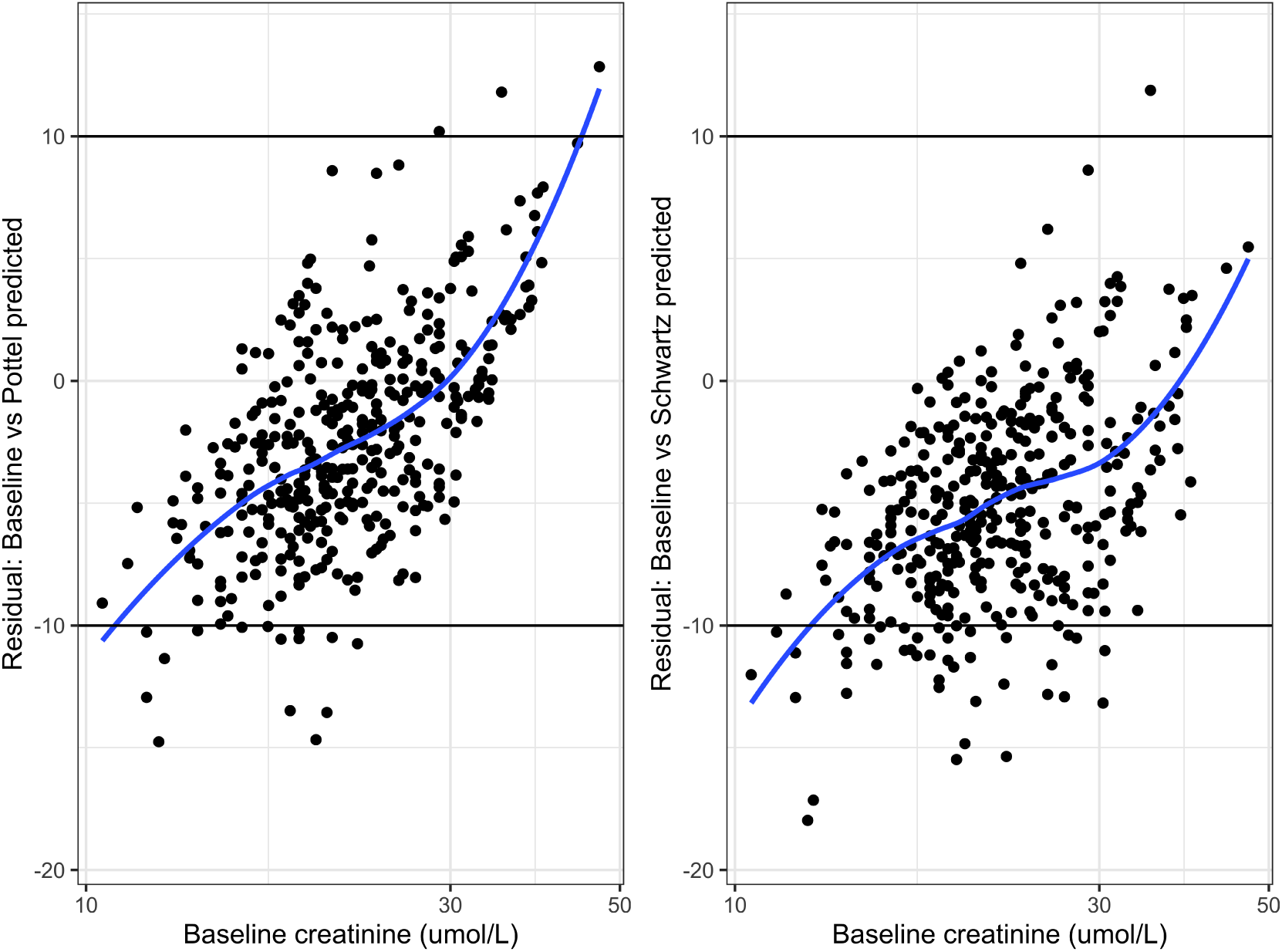
Residuals plot for the Pottel predicted baseline creatinine (left panel) and Schwartz predicted baseline creatinine (right panel). Blue lines show splines fits; horizontal lines show the +/− umol/L values.

**Figure S4:**
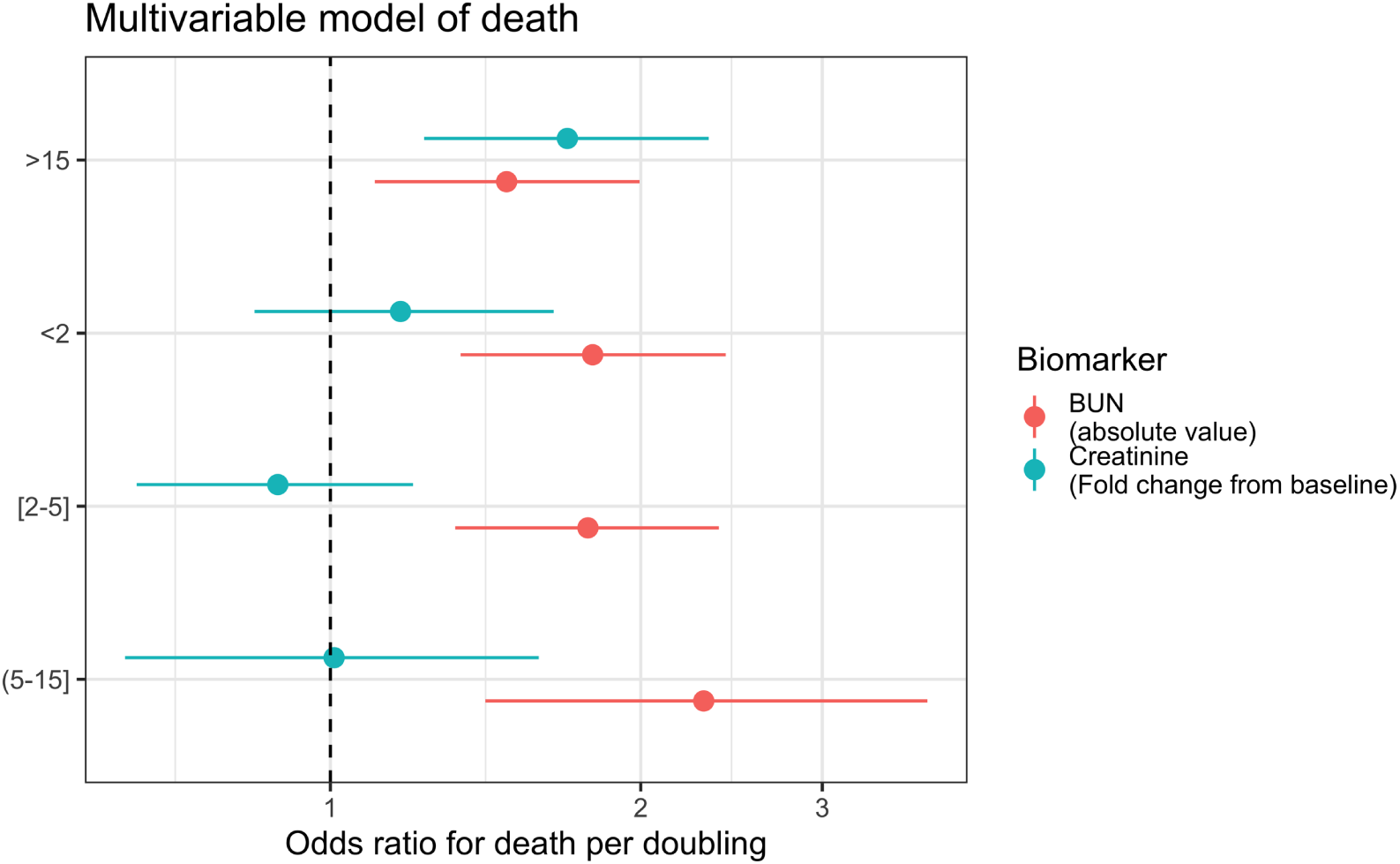
Prognostic value characterised as the odds ratio for death per doubling for the creatinine fold change versus a doubling of BUN by age group (interaction term).

**Figure S5:**
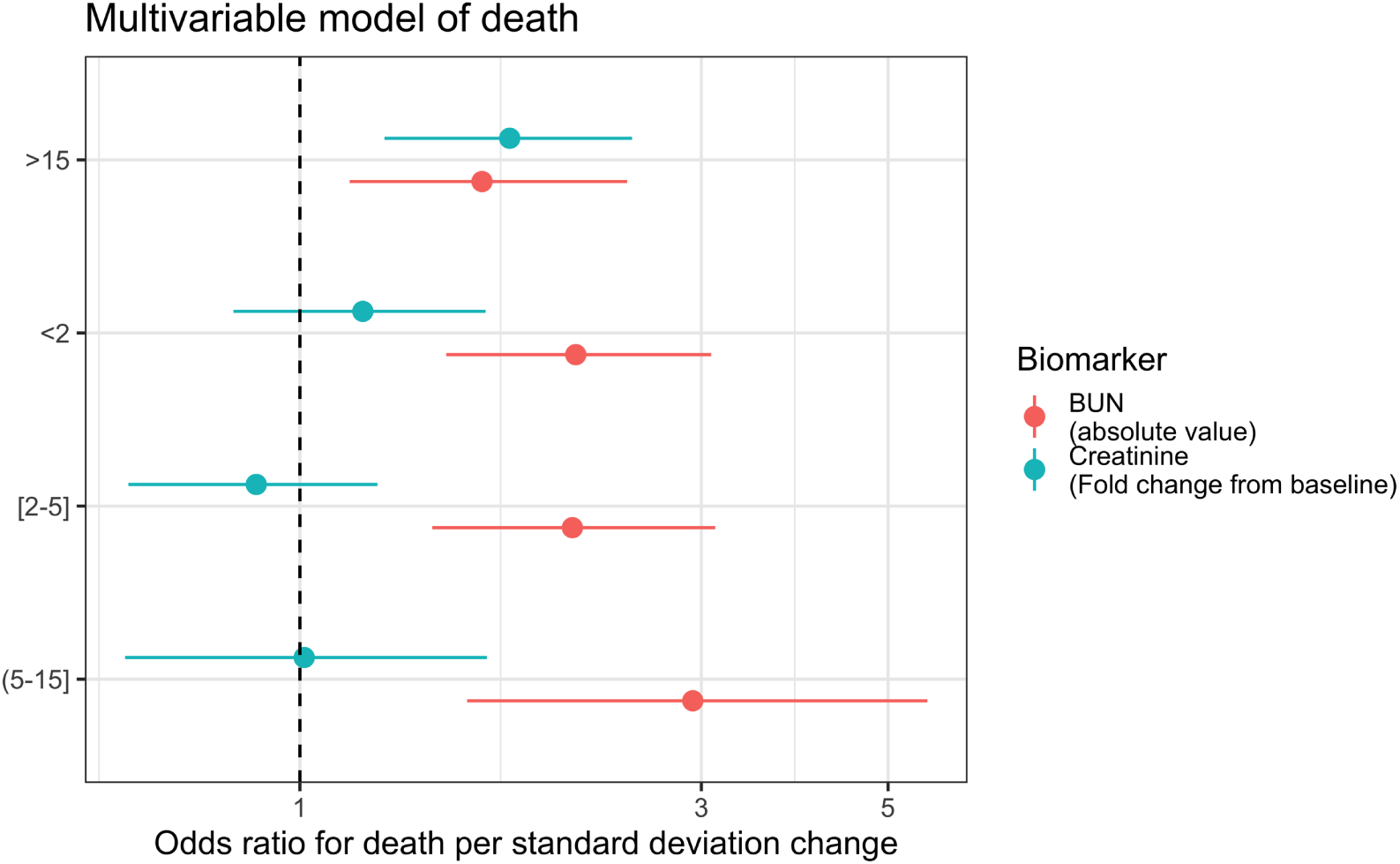
Prognostic value characterised as the odds ratio for death per standard deviation change for the log_2_ creatinine fold change versus the log_10_ BUN by age group (interaction term).

**Figure S6:**
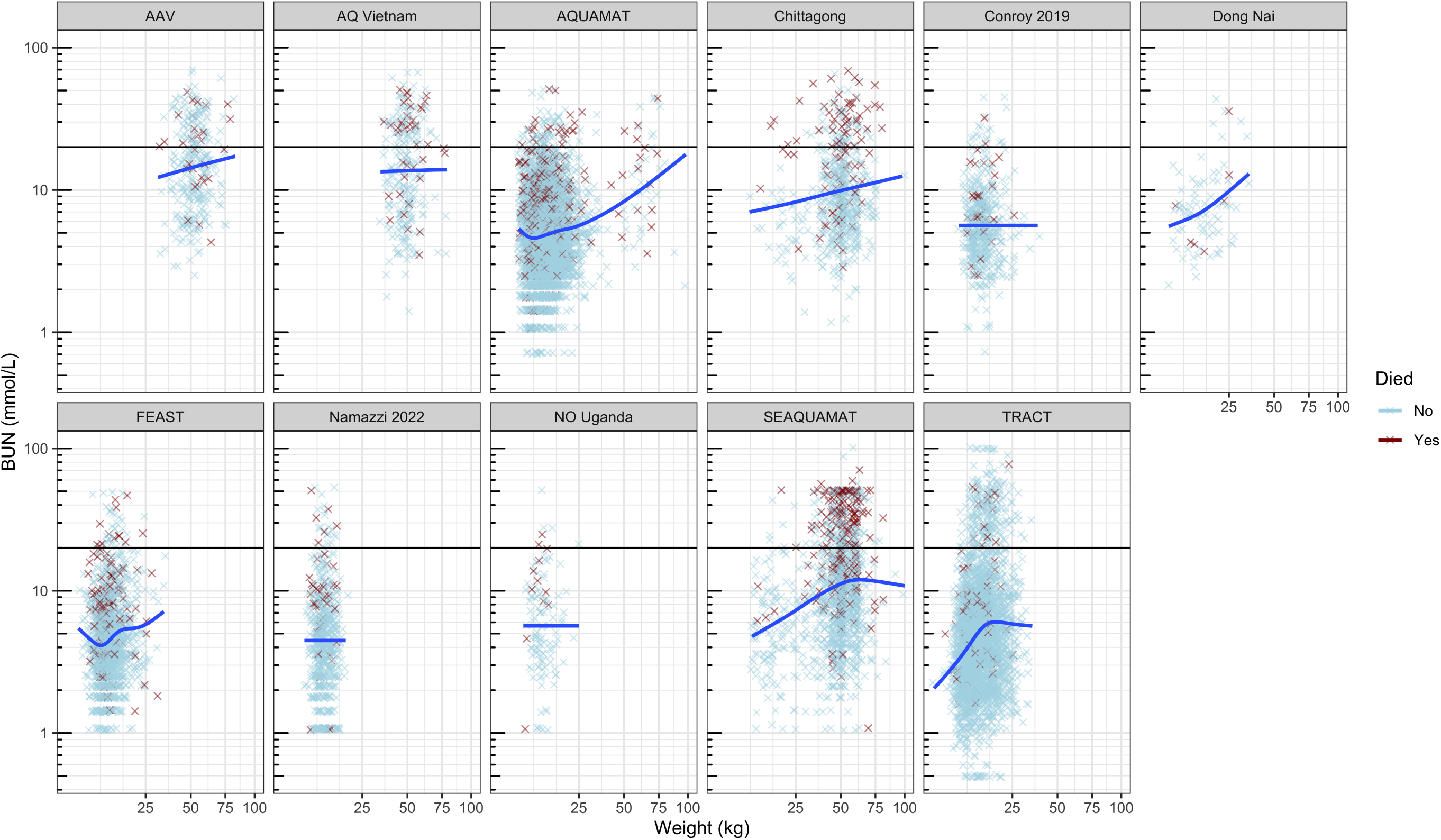
BUN (on the log_10_ scale) as a function of weight (on the sqrt scale), stratified by clinical study. Blue lines show spline fits.

**Figure S7:**
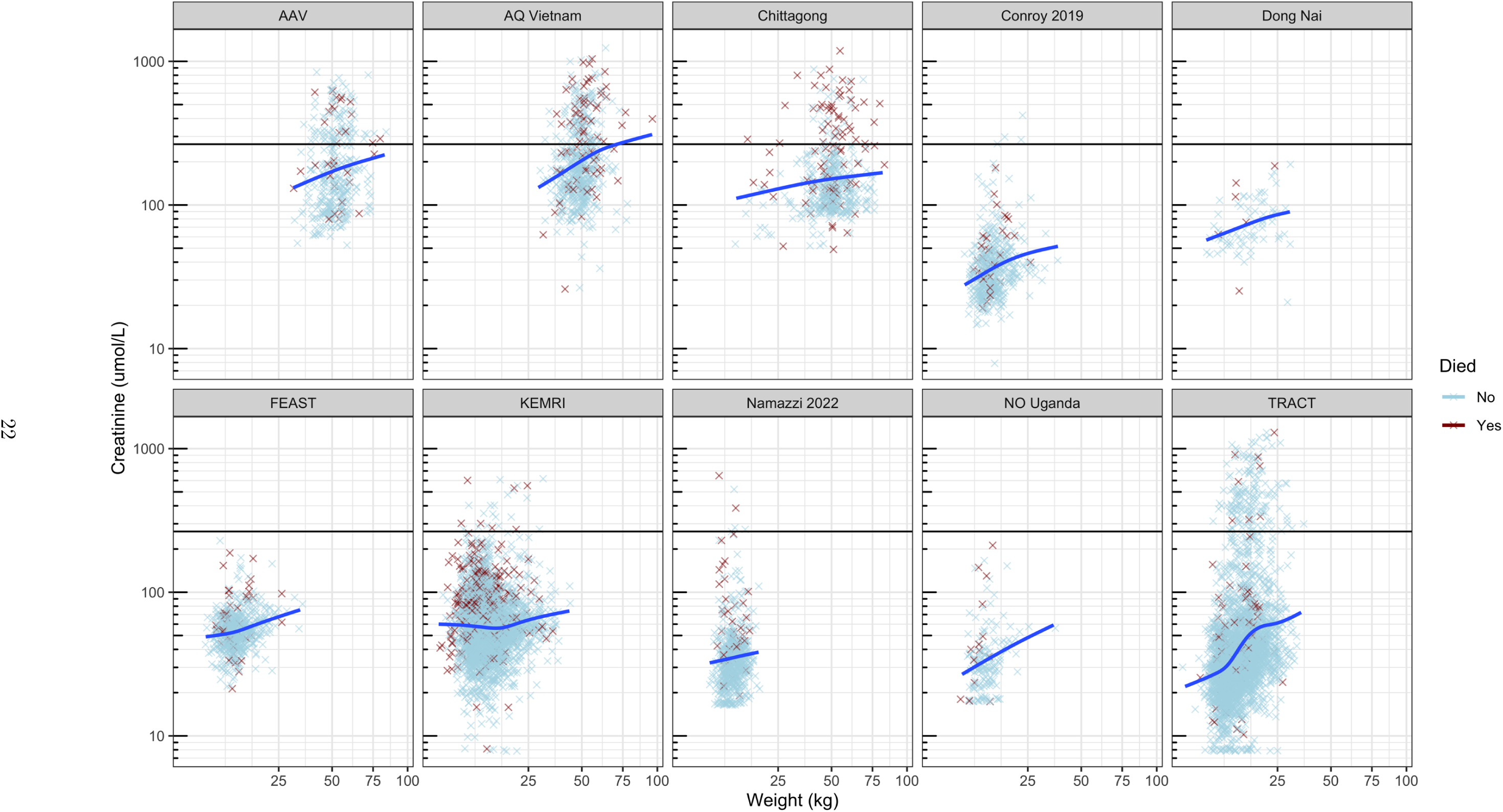
Creatinine (log_10_ scale) as a function of weight (sqrt scale), stratified by clinical study. Blue lines show spline fits.

**Figure S8:**
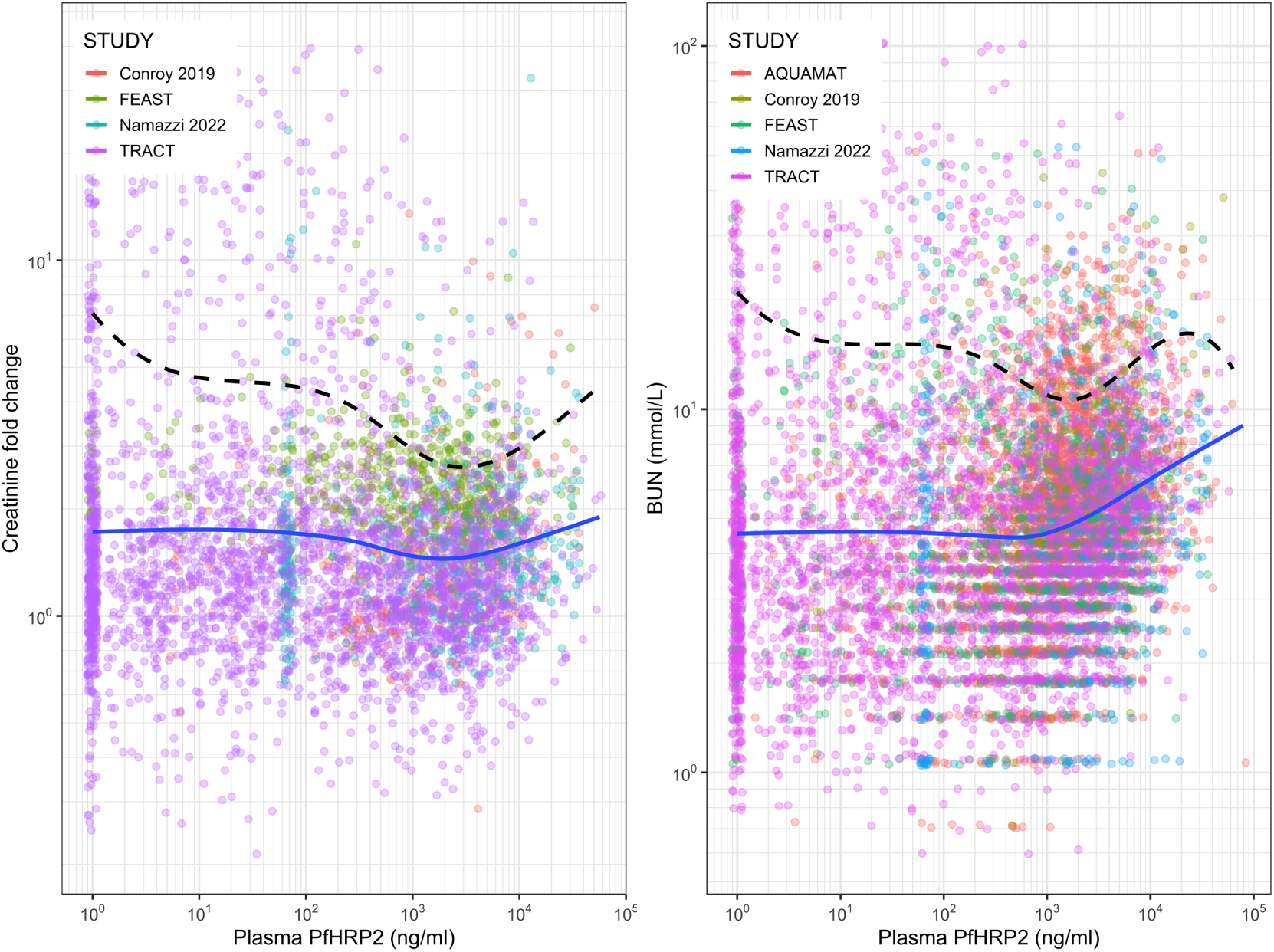
Relationship between creatinine fold change from predicted baseline (left) or absolute BUN (right) and plasma *Pf* HRP2 (a measure of parasite biomass). The blue line shows the fitted trend line for the mean value, the dashed line the quantile regression fit for the 90th percentile.

**Figure S9:**
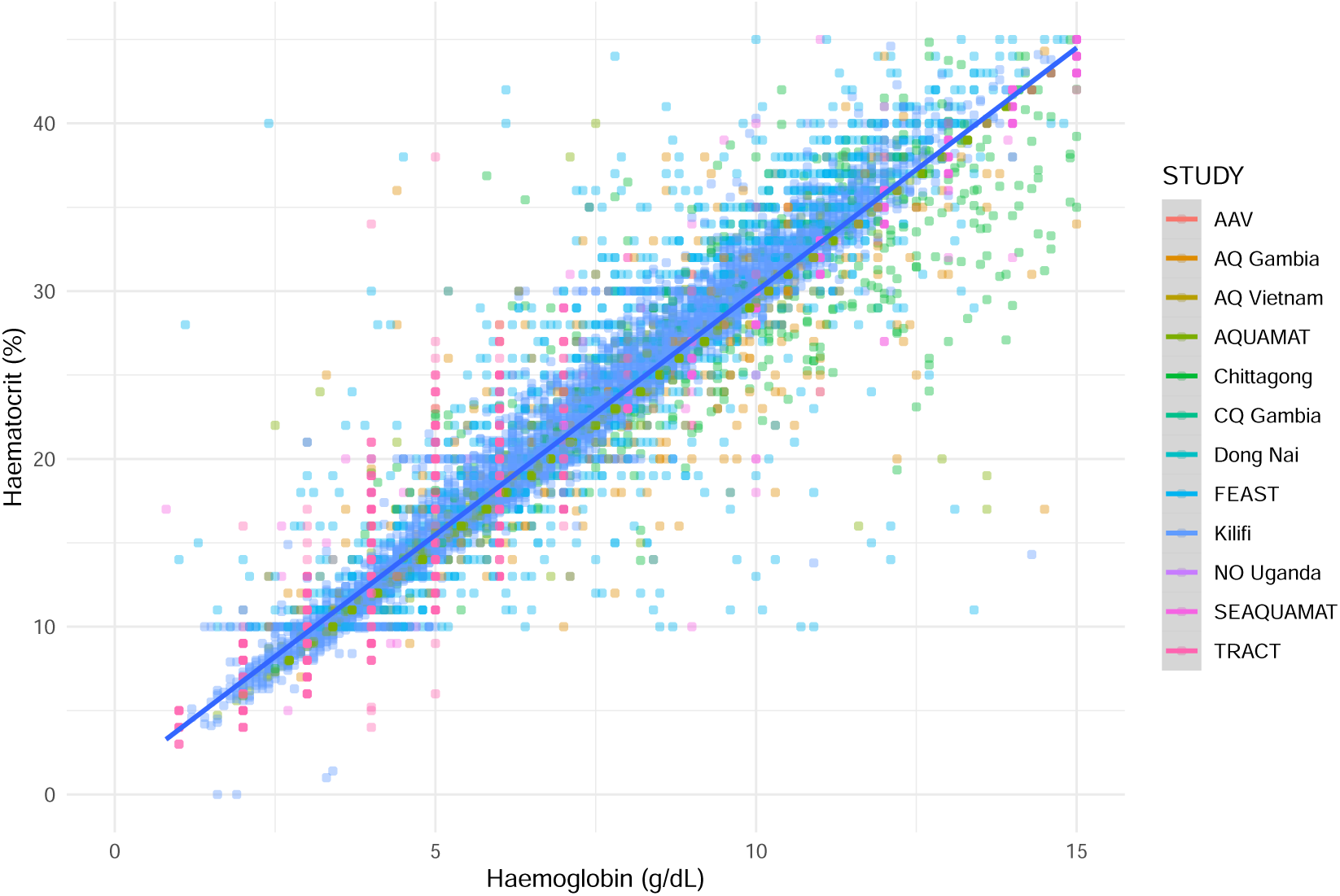
Relationship between haemoglobin (g/dL) and haematocrit on admission in 17,523 patients. The line shows the 3x transformation between the two.

## Notes

### Competing Interest Statement

The authors have declared no competing interest.

### Author Declarations

This study uses existing pseudonymised data which cannot be linked to individuals. As such, this analysis did not require additional ethical approval according to the guidelines of the Oxford Central University Research Ethics Committee.

